# Systematic identification of modifiable risk factors and drug repurposing options for Alzheimer’s disease: Mendelian randomization analyses

**DOI:** 10.1101/2020.07.14.20153726

**Authors:** Chong Wu, Lang Wu, Jingshen Wang, Lifeng Lin, Yanming Li, Qing Lu, Hong-wen Deng

## Abstract

**Introduction:** Several Mendelian randomization studies have been conducted, which identified multiple risk factors for Alzheimer’s disease (AD). However, they typically focus on a few pre-selected risk factors.

**Methods:** Two-sample Mendelian randomization (MR) study was used to systematically examine the potential causal associations of 1,054 risk factors/medical conditions and 28 drugs with the risk of late-onset AD. To correct for multiple comparisons, the false discovery rate was set at *<*0.05.

**Results:** There were strong evidence of a causal association between glioma risk, reduced trunk fat-free mass, lower education levels, lower intelligence and a higher risk of AD. For 28 investigated treatments (such as antihypertensive drugs), we found limited evidence for their associations.

**Conclusion:** MR found robust evidence of causal associations between glioma, trunk fat-free and AD. Our study also confirms that higher educational attainment and higher intelligence are associated with a reduced risk of AD.

## Introduction

Alzheimer’s disease (AD) is the leading cause of dementia and places a tremendous burden on people living with AD, their families, and caregivers^1^. The main hallmarks of AD are amyloid plaques and neurofibrillary tangles, which lead to a progressive loss of memory and cognition^2^. Apart from a few established risk factors, such as age, family history, and apolipoprotein E (APOE) e4 allele^3^, the etiology of AD is largely unknown. Furthermore, drug discovery and development for AD remain unsatisfactory, and currently, there is no cure for AD^4^. This has led to substantial interest in promoting disease prevention and treatment by targeting modifiable risk factors and exploring drug repurposing, a strategy for identifying new medical uses of existing drugs^5^.

Mendelian randomization (MR), a causal inference approach to identify risk factors and drug repurposing options, has garnered substantial interest. This is because findings from conventional observational studies are susceptible to unmeasured confounders and data from randomized controlled trials^6,7^ are scarce. MR estimates the unbiased causal effect between an exposure and a health outcome by using genetic variants, typically in the form of single-nucleotide polymorphisms (SNPs), as instruments^8,9^. Because genetic variants are randomly assorted from parents and are fixed at conception, MR can be conceptualized as a “genetic randomized controlled trial” and hence is less likely to be affected by unmeasured confounders and reverse causation that could bias the associations^8,9^. Furthermore, MR can be performed by using summary statistics from genome-wide association studies (GWAS), in an approach known as two-sample MR^10^. Specifically, in two-sample MR, the SNP-exposure and SNP-outcome associations are obtained from two independent GWAS summary data, which are easier to obtain and generally have larger sample sizes. By leveraging existing GWAS data, two-sample MR substantially increases the statistical power as well as the number of exposures that could be investigated.

Several MR analyses have been conducted for AD, which identified multiple risk factors, such as educational attainment^11^, LDL cholesterol level^12^, systolic blood pressure^13^, alcohol consumption^14^, and smoking quantity^13^. However, they typically focus on a few pre-selected risk factors, and the results may be over-optimistic due to publication bias. Importantly, recently it has been proposed that MR could be applied to predict promising drug repurposing options. For AD, previous work only investigated a few medications such as antihypertensive drugs^15^ for drug repurposing options. To our knowledge, no promising drug candidates have been found through MR analyses. To decipher potentially causal risk factors and investigate potential drug repurposing options, we applied two-sample MR to systematically examine the potential causal associations of 1,054 risk factors and 28 drugs with AD.

## Methods

### Risk factors and potential drug repurposing options

We obtained GWAS summary data for risk factors/medical conditions and medications/treatments from the IEU GWAS database. To obtain robust estimation, we focused on the exposures (including 1,054 risk factors/medical conditions and 28 medications/treatments) that had at least three independent genome-wide significant instrument SNPs. Details on the exposures are presented in Supplementary Tables 1 and 2.

**Table 1.** Odds ratios for associations between significant risk factors/medical conditions and Alzheimer’s disease risk. OR=odds ratio; 95% CI=95% confidence interval. IVW represents the widely- used inverse variance weighted method, weighted median MR is a robust MR method that provides a consistent estimator when up to 50% of the instrumental variables are invalid. MR-Egger intercept test and MR-PRESSO global test are used to check directional pleiotropy and heterogeneity.

| Exposure | IVW |  |  | Weighted median |  |  | MR-Egger Intercept | MR-PRESSO global |
| --- | --- | --- | --- | --- | --- | --- | --- | --- |
|  | OR | 95% CI | *P* | OR | 95% CI | *P* | *P* | *P* |
| **College or University degree** | 0.41 | 0.28-0.60 | $4.0 \times 10^{-6}$ | 0.38 | 0.23-0.63 | $1.6 \times 10^{-4}$ | 0.39 | <0.001 |
| **Years of schooling** | 0.68 | 0.58-0.80 | $5.0 \times 10^{-5}$ | 0.62 | 0.49-0.77 | $2.8 \times 10^{-5}$ | 0.22 | 0.02 |
| **A levels/AS levels or equivalent** | 0.27 | 0.15-0.50 | $2.5 \times 10^{-5}$ | 0.35 | 0.14-0.87 | 0.023 | 0.48 | 0.42 |
| **Trunk fat-free mass** | 0.78 | 0.68-0.89 | $2.0 \times 10^{-4}$ | 0.76 | 0.62-0.93 | $7.2 \times 10^{-3}$ | 0.93 | 0.01 |
| **Intelligence** | 0.72 | 0.60-0.86 | $4.6 \times 10^{-4}$ | 0.66 | 0.52-0.83 | $3.9 \times 10^{-4}$ | 0.94 | <0.001 |
| **Glioma** | 1.13 | 1.06-1.21 | $4.8 \times 10^{-4}$ | 1.11 | 1.02-1.21 | 0.015 | 0.40 | - |
| **Qualifications: None of the above** | 3.68 | 1.77-7.67 | $5.0 \times 10^{-4}$ | 3.8 | 1.39-10.36 | $9.2 \times 10^{-3}$ | 0.22 | 0.09 |

**Table 2.** Odds ratios for associations between suggestive risk factors/medical conditions and Alzheimer’s disease. OR=odds ratio; 95% CI=95% confidence interval. IVW represents the widely-used inverse variance weighted method, weighted median MR is a robust MR method that provides a consistent estimator when up to 50% of the instrumental variables are invalid. MR-Egger intercept test and MR-PRESSO global test are used to check directional pleiotropy and heterogeneity.

| Exposure | IVW |  |  | Weighted median |  |  | MR-Egger Intercept | MR-PRESSO global |
| --- | --- | --- | --- | --- | --- | --- | --- | --- |
|  | OR | 95% CI | *P* | OR | 95% CI | *P* | *P* | *P* |
| **Neoplasms** | 0.06 | 0.01-0.30 | $6.2 \times 10^{-4}$ | 0.04 | 0.01-0.34 | $3.0 \times 10^{-3}$ | 0.16 | 0.63 |
| **Basal metabolic rate** | 0.78 | 0.68-0.91 | $9.4 \times 10^{-4}$ | 0.78 | 0.64-0.96 | $1.8 \times 10^{-2}$ | 0.90 | <0.001 |
| **Monocyte count** | 0.87 | 0.80-0.94 | $1.1 \times 10^{-3}$ | 0.87 | 0.76-1.00 | $4.5 \times 10^{-2}$ | 0.82 | <0.001 |
| **Vitamin D levels** | 0.6 | 0.44-0.82 | $1.3 \times 10^{-3}$ | 0.65 | 0.46-0.92 | $1.4 \times 10^{-2}$ | 0.35 | 0.47 |
| **Height** | 0.90 | 0.84-0.96 | $1.8 \times 10^{-3}$ | 0.90 | 0.82-0.99 | $3.8 \times 10^{-2}$ | 0.61 | 0.002 |
| **Job involves mainly walking or standing** | 1.89 | 1.24-2.89 | $3.2 \times 10^{-3}$ | 1.87 | 1.04-3.39 | $3.7 \times 10^{-2}$ | 0.33 | 0.38 |
| **Standing height** | 0.87 | 0.79-0.96 | $4.4 \times 10^{-3}$ | 0.86 | 0.76-0.98 | $2.5 \times 10^{-2}$ | 0.89 | <0.001 |

### Late-onset AD

We obtained GWAS summary data (i.e., effect size estimates and their standard errors) for the associations between the genetic variants and late-onset AD status from the International Genomics of Alzheimer’s Project (IGAP), which has been described elsewhere^16^. In brief, this study includes data from 17,008 late-onset AD cases and 37,154 cognitively normal elderly controls of European ancestry in four cohorts, including the Alzheimer’s Disease Genetics Consortium (ADGC), the Genetic and Environmental Risk in Alzheimer’s Disease (GERAD) Consortium, the European Alzheimer’s Disease Initiative (EADI), and the Cohorts for Heart and Aging Research in Genomic Epidemiology (CHARGE) Consortium. The average age of participants was 71 years. The associations of late-onset AD with genetic variants were analyzed by a logistic regression model adjusting for covariates of age, sex, and genetic principal components. Since allele frequencies in the IGAP data were not reported, we used allele frequencies from 503 European samples in 1000 Genomes phase 3.

### Genetic instrumental variables

For each exposure, we selected genetic variants (i.e., SNPs) associated with the exposure at the genome-wide significance threshold (*P* < 5 × 10^−10^). We selected independent genetic variants—that is, no linkage disequilibrium (*r*^*^ < 0.001 with an extension of 10000 Kb in the European reference panel of 1000 Genomes phase 3) with other selected genetic variants. When genetic variants were in linkage disequilibrium, we chose the variant with the lowest *P* value. For genetic variants that were not available in the outcome dataset (i.e., IGAP), we used proxies (*r*^*^ > 0.8 in the European reference panel of 1000 Genomes phase 3) where available. We further harmonized exposure-outcome datasets by the R package “twosampleMR”^17^, allowing the strand direction of ambiguous SNPs to be inferred by leveraging allele frequency information.

### Mendelian randomization analysis

We applied the widely-used inverse variance weighted (IVW) method^18^ to estimate the overall causal effect of an exposure on AD risk. IVW aggregates causal ratio estimates provided by each genetic instrument variant and yields a consistent and efficient estimator when all used genetic variants are valid instrumental variables. Valid instrumental variables need to meet three key assumptions (Figure 1): 1) they are associated with the exposure; 2) they are not associated with any confounders; and 3) they can affect the outcome through the exposure only—that is, no horizontal pleiotropy.

**Figure 1.**
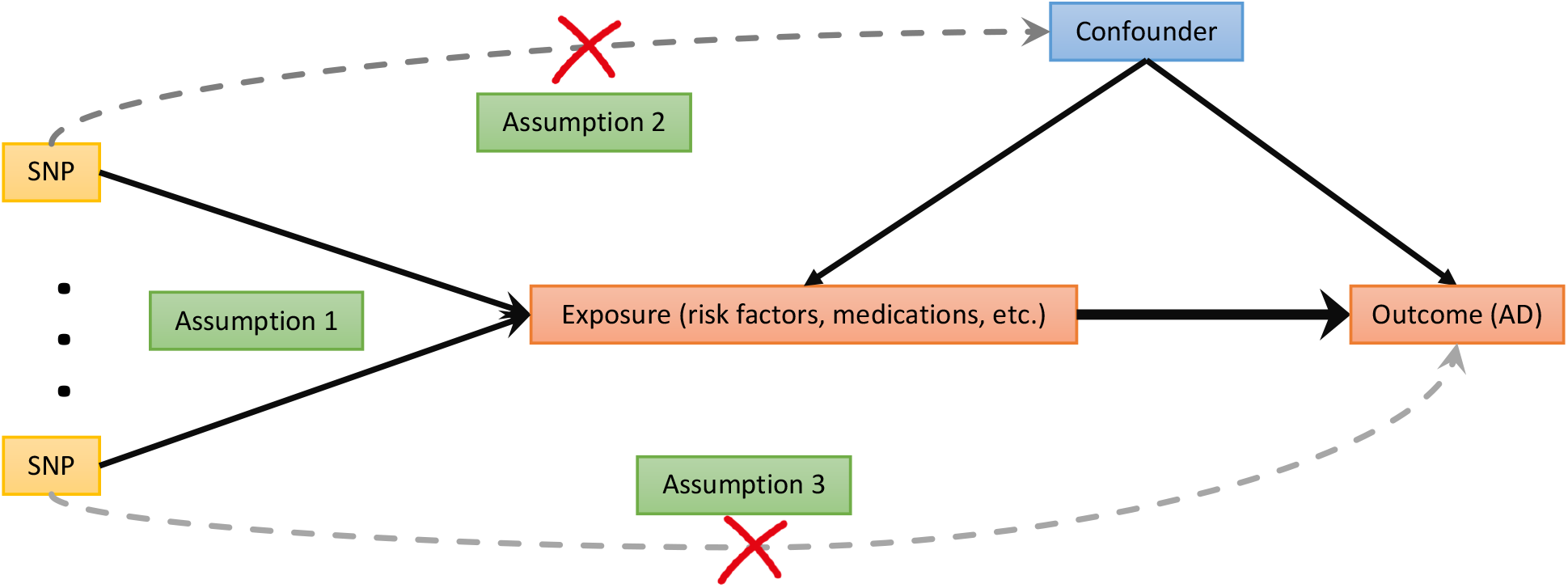
Principles of Mendelian randomization analysis and assumptions that need to be met to obtain unbiased estimates of causal effects. Genetic variants have effect on the exposure (Assumption 1) and are not associated with confounders (Assumption 2) or the outcome directly (Assumption 3).

To obtain valid results and account for the potential violations of valid instrumental variable assumptions, we conducted sensitive analyses by applying several methods that are robust to horizontal pleiotropy at the cost of reduced statistical power. These methods include weighted median MR^19^, MR-Egger regression^20^, and Mendelian randomization pleiotropy residual sum and outlier (MR-PRESSO) test^21^. Weighted median MR provides a consistent estimator when up to 50% of the instrumental variables are invalid. MR-Egger regression allows all the genetic variants to be invalid instrumental variables, and it is equivalent to a weighted regression with the slope representing the causal estimate and the intercept representing the bias due to horizontal pleiotropy. MR-PRESSO detects horizontal pleiotropy via a global test, corrects for horizontal pleiotropy via outlier removal, and tests if the causal estimates are significantly different before and after outlier correction.

Results are presented as odds ratios (ORs) per genetically predicted increase in each exposure (i.e., modifiable risk factors or drug repurposing options) with default scale reported in each GWAS. For the binary risk factors, the estimates represent the OR per 1 unit higher log odds of the exposure. To correct for multiple comparisons, the significance level was set to 5 × 10^−4^, a threshold corresponding to a false discovery rate (FDR) < 0.05. Moreover, 5 × 10^−4^ < P < 5 × 10^−3^ was used to determine suggestive associations. The associations with P > 0.05 in weighted median MR analysis were considered to be spurious. All statistical analyses were conducted using R version 3.6.1 (R foundation).

## Results

We firstly present the medical conditions and modifiable risk factors with robust causal effects on AD (Table 1). Supplementary Table 3 includes the estimates generated from the IVW, weighted median analysis, and MR-Egger, as well as the numbers of instrument variants used for each risk factor/medical condition we investigate. We also present suggestive associations (Table 2) and the results for drug repurposing options.

### Glioma

Genetically predicted glioma status was significantly associated with a higher risk of AD. The OR was 1.13 (95% confidence interval [CI]: 1.06 to 1.21; P = 4.8 × 10^−4^) per unit higher log odds of having glioma. By using the MR-Egger Intercept test, we found limited evidence of horizontal pleiotropy (P = 0.40). Also, the association was consistent in sensitivity analysis as the weighted median provided similar conclusions but with less precision. Leave one out analysis also indicated that no single SNP had an influential influence on the estimation (Supplementary Figure 1). In bi-directional MR analysis, we found no evidence that genetically predicted AD was associated with glioma (P = 0.55).

### Trunk fat-free mass

Genetically predicted trunk fat-free mass was significantly associated with a reduced risk of AD. The OR was 0.78 (95% CI: 0.68 to 0.89; P = 2.0 × 10^−4^) per standard deviation (5.17 Kg) higher of fat-free mass. There was limited evidence of directional pleiotropy or heterogeneity based on the MR-Egger intercept test (P = 0.94) and MR-PRESSO global test (P = 0.01). The association remained robust in the weighted median MR and leave one out analyses (Supplementary Figure 2).

### Education and intelligence

Genetically predicted higher education was significantly associated with a reduced risk of AD. The ORs were 0.68 (95% CI: 0.58 to 0.80; P = 5 1 × 10^−6^) per year of education completed and 0.41 (95% CI: 0.28 to 0.60; P = 4 0 × 10^−6^) per unit higher log odds of having completed college or university, 0.27 (95% CI: 0.15 to 0.50; P = 2 5 × 10^−5^) per unit higher log odds of having completed A levels/AS levels and equivalent, and 3.68 (95% CI: 1.77 to 7.67; P = 5 0 × 10^−4^) per unit higher log odds of not having education. Not having education means participants select “None of the above” for the question: “which of the following qualifications do you have? (You can select more than one)” Possible answers were: ‘college or university degree/A levels or AS levels or equivalent/O levels or GCSE or equivalent/CSEs or equivalent/NVQ or HND or HNC or equivalent/Other professional qualifications, for example, nursing, teaching/none of the above/prefer not to answer’. Also, there was a significant association between genetically predicted intelligence and lower odds of AD (OR: 0.72; 95% CI: 0.60 to 0.86; P = 4 6 × 10^−4^). MR-Egger analyses showed no evidence of directional pleiotropy. While completed college or university and intelligence had heterogeneity problems as indicated by MR-PRESSO global test, the associations remained similar after removing outliers (Supplementary Table 4). There was no outlier detected for either completing A levels/AS levels and equivalent or years of schooling. Furthermore, the associations were consistent in sensitive analyses as the weighted median method yielded similar (yet less precise) results.

### Suggestive associations

We found several suggestive associations. Specifically, we found suggestive evidence of an inverse association between neoplasms and AD. There was a suggestive association between genetically predicted higher basal metabolic rate and lower odds of AD (OR: 0.78; 95% CI: 0.68 to 0.91; P = 6 2 × 10^−4^). Genetically predicted monocyte count showed suggestive association with a reduced risk of AD. We found suggestive evidence for a positive association between AD and having a job that involves mainly walking or standing.

We also found a suggestive association between higher height and a reduced risk of AD (OR: 0.90; 95% CI: 0.84 to 0.96; P = 1 8 × 10^−3^). Genetically predicted vitamin D levels were associated with lower odds of AD suggestively. The OR was 0.60 (95% CI: 0.44 to 0.82; P = 1 3 × 10^’0^). These associations remained robust in the weighted median MR and there was limited evidence of pleiotropy and heterogeneity.

### Medications/treatments

There was limited evidence for an overall effect of 28 medications/treatments we investigated on the risk of AD, as most estimates provided limited evidence to exclude the possibility of no association (Supplementary Table 5). For example, antihypertensive drugs amlodipine and atenolol had an OR of 0.47 (95% CI: 0.05 to 4.49; P = 0.51) and 0.14 (95% CI: 0 to 6.28; P = 0.31), respectively.

## Discussion

Unlike existing studies, this is the first Mendelian randomization (MR) study that systemically examined the causal associations of 1,054 risk factors and 28 drugs with AD. This MR analysis found strong evidence for a positive association between genetically predicted glioma and risk of AD, and an inverse association between genetically predicted trunk fat-free mass and risk of AD. We confirmed that a higher educational attainment and a higher intelligence are associated with a reduced risk of AD.

### Comparison with existing literature

Conventional observational studies provide inconsistent evidence for the associations between glioma and AD: some suggest an inverse association between the two^22^, while others suggest a positive association^23^. These inconsistent findings may result from unadjusted confounding factors. Through MR analyses, we found strong evidence to support that glioma was significantly associated with a higher risk of AD. This finding is also consistent with a recent study ^23^. By using the bidirectional MR analysis, we did not detect an association between genetically predicted AD and glioma, further supporting that glioma increases the risk of AD and not the other way around.

We found that genetically predicted trunk fat-free mass was significantly associated with lower risk of AD. This is in line with the results from observational studies showing AD patients had lower lean tissue mass than healthy elderly individuals^24^.

The evidence from observational studies and previous MR analyses consistently supports that genetically predicted high educational attainment^11^ to be associated with a reduced risk of AD. Furthermore, with a larger GWAS of 78,398 individuals for intelligence^25^, we were able to use an increased number of instrumental variables to 14. With such improvement, we confirmed that higher intelligence was significantly associated with reduced risk of AD, which was only a suggestive association in the earlier MR analyses^11^.

For the 28 drug repurposing options we analyzed, we did not observe evidence supporting their potential effects on reducing AD risk. Specifically, consistent with previous MR analyses^15^, there was limited evidence to support the effects of antihypertensive drugs on AD. For cholesterol drugs (such as ezetimibe, rosuvastatin, simvastatin, and atorvastatin), anti-diabetic drugs (such as gliclazide and metformin), and anti-inflammatory drugs (such as ibuprofen and aspirin), we also did not find any evidence supporting their effects on reducing AD risk.

### Strengths and limitations

The strengths of this study include the two-sample MR design and a systematically assessment of risk factors, medical conditions, and drug repurposing options in relation to AD. By leveraging large-scale GWAS data, the two-sample MR design simultaneously reduces the risk of confounding and improves the statistical power; by systematically accounting for a large number of potentially exposures, it also reduces the concern of publication bias.

Causal inference of the associations identified in this study, however, relies on several assumptions (Figure 1) that may be violated in the analyses. First, because the same GWAS data have been used to select instruments and to estimate SNP-exposure associations, our findings might be affected by weak instrument bias. Nevertheless, given the nature of the two-sample MR analysis, the weak instruments only induce a bias towards the null hypothesis. Hence, such a bias will not inflate the Type 1 error rate for false positive findings. Second, this study cannot completely rule out the risk of horizontal pleiotropy, which is challenging for all MR analyses. We addressed this by conducting several sensitive analyses, including the weighted median, MR-Egger, MR-PRESSO global test, and leave one out analysis. Inconsistent results from the IVW and sensitive analyses were treated as spurious. Third, because of the limited sample size of some GWAS data, the power was limited for some exposures, and thus the null findings may result from the Type 2 error with false negative findings.

Another limitation relating to drug repurposing discovery is that drugs/treatments typically are given for short periods, whereas MR estimates the effect of lifelong exposures. This inconsistency indicates that the effect sizes estimated in this study will likely be different from those observed in clinical trials or clinical practice. There are two new GWAS for AD^26,27^, but we were unable to use them due to the data usage requirements and overlapping samples for many exposures; instead, data from IGAP were used in this article. Finally, the current analyses were focused on individuals of European ancestry; special attention is needed when generalizing our findings to other populations.

## Conclusions

In summary, by using the MR design, we observe evidence that having glioma is associated with a higher risk of AD and higher trunk fat-free mass is associated with a reduced risk of AD. Our study also confirms that a higher educational attainment and a higher intelligence are associated with a reduced risk of AD.

## Data Availability

The proposed study uses publically available GWAS datasets (https://gwas.mrcieu.ac.uk).

## Contributors

CW analyzed the data, drew the figures, and wrote the first draft of the manuscript. CW is the guarantor of this work. All authors contributed to the interpretation of the results and critical revision of the manuscript for important intellectual content and approved the final version of the manuscript.

## Funding

This study is supported by the Committee on Faculty Research Support (COFRS) grant at Florida State University.

## Competing interests

No potential conflicts of interest were disclosed by the authors.

## Acknowledgments

We thank the individuals involved in the GWAS datasets for their participation and the research teams for their work on collecting, processing, and sharing these datasets.

## Notes

### Competing Interest Statement

The authors have declared no competing interest.

### Funding Statement

No external funding was received.

### Author Declarations

No IRB/oversight is required as the proposed study only uses summarized data.

